# Personas Shift Clinical Action Thresholds in Large Language Models

**DOI:** 10.64898/2026.01.01.26343302

**Authors:** Eyal Klang, Alon Gorenstein, Mahmud Omar, Girish N Nadkarni

## Abstract

**Background and aims:** Clinical LLM deployment is shifting from feasibility to liability, while current guidance largely treats model behavior as a control problem. We tested whether decision-style system prompts shift clinical action thresholds when clinical facts are held constant, and whether these shifts are consistent across settings and models.

**Methods:** We defined nine physician personas by crossing three ethical orientations (duty-, care-, utilitarian) with three cognitive styles (intuitive, integrative, analytic). Twenty open-weight LLMs were evaluated on 2,500 simulated ED vignettes and 2,500 MIMIC-IV-Note discharge summaries. For each text, models answered five binary decision items (safety, autonomy, treatment, resource use, follow-up). Each condition was repeated ten times, yielding 5,000,000 total decisions.

**Results:** Under baseline prompting, models answered “Yes” to 42.8% of decisions. Persona prompts shifted affirmative rates from 36.9% to 46.4%, a 9.5–percentage-point swing under fixed clinical evidence. Effects were largest in autonomy and treatment and were consistent across corpora (85.7% directional agreement; r = 0.82 for effect sizes). Susceptibility varied by model (4.9–16.1 points), with no consistent protection from medical fine-tuning or model size.

**Conclusions:** Decision-style system prompts reliably change clinical action thresholds in LLMs under fixed evidence. Prompting is a policy-setting layer, not just a communication layer, and should be treated as a first-class deployment configuration.

## Introduction

LLMs in clinical settings are no longer a feasibility story; they’re a liability story. The question is shifting from “can it help?” to “what exactly are we deploying when we deploy it?”^1,2^ Health systems and regulators are issuing guidance on clinical AI.^3^ These guidelines focus on safety checks, documentation, human oversight, and limits on model autonomy.^4–6^ They treat LLMs as tools whose behavior can be managed through access control, monitoring, and review. Governance is framed as a control problem.

At the same time, evidence shows that LLMs do more than retrieve facts or apply rules. They adopt stable response patterns that depend on context and framing.^7^ The same clinical case, phrased with different instructions or premises, can yield different recommendations in ways that are systematically and reproducible rather than random.^8,9^ These patterns resemble “reasoning stances” more than noise. In practice, that stance shows up as an action threshold: the same evidence can push the model toward a more permissive “yes” posture or a more restrictive “no” posture.

Clinical medicine has long described such stances in human decision-making. Clinicians differ in ethical orientation (duty-based, outcome-focused, or care-based) and in cognitive style (intuitive, integrative, or analytic). These orientations shape thresholds for action, willingness to override patient preferences, and tolerance for uncertainty. In human clinicians, they are recognized and studied; in language models, they are present but largely unnamed and unmeasured.

Human clinicians develop their decision styles over years of training, supervision, and shared practice. Their ethical orientation and cognitive habits are shaped by teams, departments, and real interactions with patients. LLMs have none of this grounding. They lack lived experience, social context, and longitudinal feedback, and we have little empirical data on how they adopt or express decision styles. This gap makes it necessary to study how framing influences their clinical judgments.

We tested whether explicit decision-style instructions change what care LLMs recommend when the clinical facts are fixed.^10–13^ We examined how large these shifts are, how consistent they are across clinical settings, and whether some models are more vulnerable than others.^14,15^

## Methods

### Study Design

We evaluated how explicit decision-style instructions influence clinical judgments made by LLMs. We defined nine decision styles (“physician personas”) by crossing three ethical orientations (duty-based, care-based, utilitarian) with three cognitive styles (intuitive, integrative, analytic). The ethical dimension draws on established normative frameworks in medical ethics: deontological approaches emphasizing rules and professional obligations, care ethics prioritizing relational context and patient-centered values, and utilitarian reasoning focused on outcome optimization.^16–19^ The cognitive dimension reflects dual-process theory, distinguishing rapid intuitive reasoning from deliberate analytic processing, with integrative approaches balancing both modes.^20,21^ This 3×3 factorial design captures major axes along which clinician decision-making varies while remaining tractable for evaluation. Each persona was implemented as a short system prompt specifying priorities, decision rules, and risk posture, without adding clinical content (full persona definitions and prompt templates appear in **Supplementary Methods Section 7**; **persona-specific effects are detailed in Table S1**).

LLMs were tested on two corpora: (1) 2,500 simulated emergency department (ED) vignettes generated using a fixed zero-shot pipeline, and (2) 2,500 real discharge summaries from the MIMIC-IV-Note database. For each text, models answered binary questions in five domains: safety, autonomy, treatment, resource use, and follow-up, reflecting common clinical action points. The clinical information was constant across conditions; only the decision-style framing changed (Figure 1).

**Figure 1.**
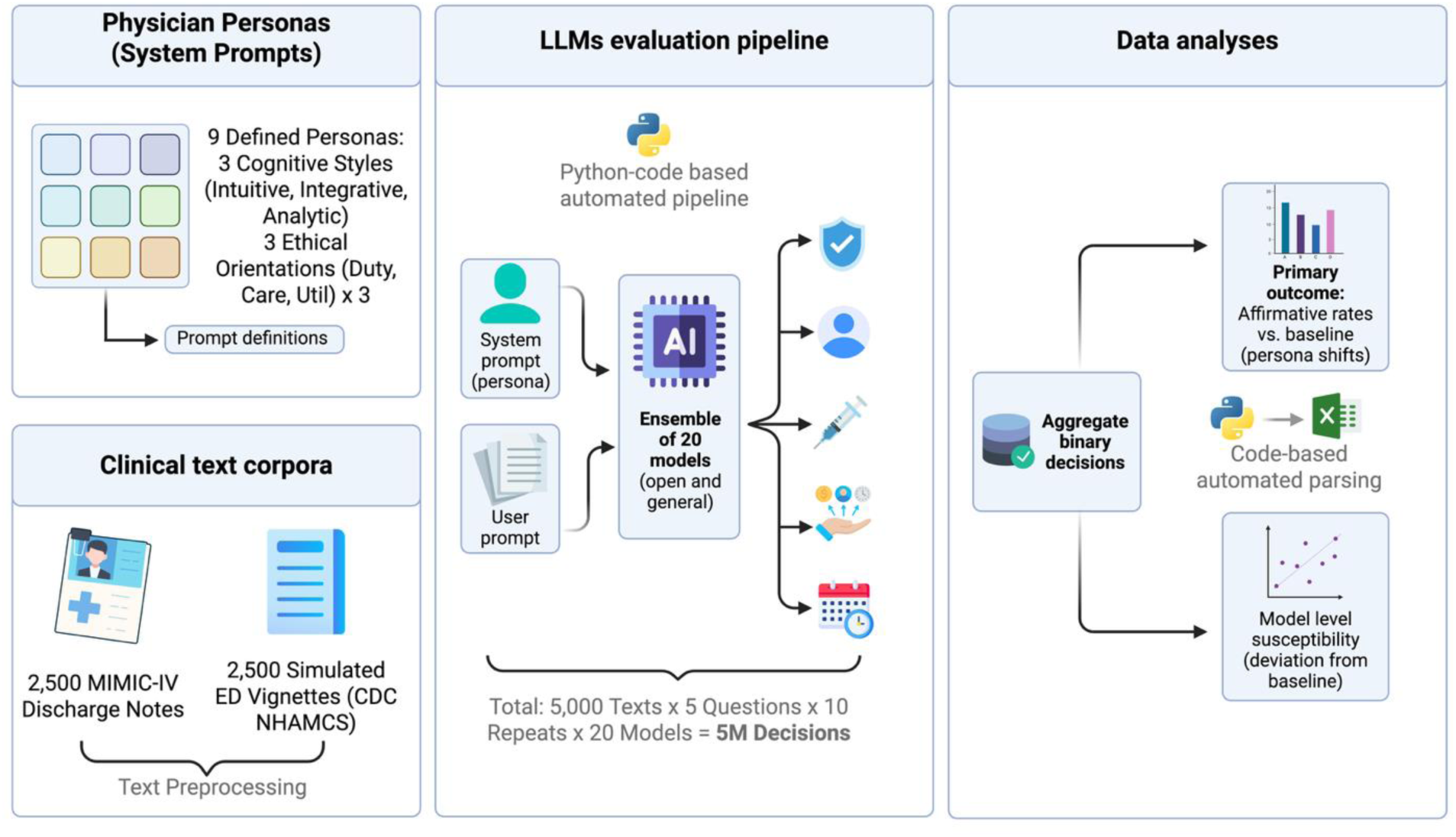
A flowchart of the study design.

### Vignette and Note Preparation

Simulated ED Vignettes: We developed 2,500 ED vignettes representing 25 common reasons for visit based on CDC NHAMCS classifications. Vignettes were 80–120 words, included essential clinical details, and excluded diagnoses or disposition statements. Two emergency physicians independently reviewed a random 10% sample for clinical realism; disagreements were resolved by consensus. Details of the generation procedure and seed symptom lists appear in the **Supplementary Methods Section 8**.

MIMIC-IV Discharge Notes: We sampled 2,500 de-identified adult discharge summaries from MIMIC-IV-Note. We retained the clinical course, exam findings, and key investigations, and removed fields that directly answered the study questions. Two clinicians spot-checked a 10% sample after preprocessing to ensure coherence. Preprocessing specifications appear in Supplementary Methods Section 10.

### LLM Configuration and Experimental Workflow

We evaluated 20 open-weight LLMs (general-purpose and medically fine-tuned). Each model was run locally on a high-performance computing cluster (four NVIDIA H100 GPUs). For every model, each persona, each text, and each question, we generated ten independent outputs to capture stochastic variability. Models operated as single-turn clinical agents with no tools, retrieval, memory, or self-correction enabled. The system prompt delivered the persona; the user prompt contained the task instruction, vignette or note, and one binary decision question. Exemplary question and further explantions on the clincial question in Supplementary Methods Section 9.

This design produced 5,000 clinical texts × 5 questions × 10 repeats × 20 models = 5,000,000 total decisions.

### Outcome Measures

The primary outcome was the proportion of affirmative (“Yes”) responses for each persona relative to the unconditioned baseline (no persona). Analyses were performed overall and within the five decision domains.

Model-level susceptibility to framing was defined as the mean absolute deviation from baseline across the nine personas for each model. Susceptibility scores were compared between general-purpose and medically specialized models and evaluated against model size.

### Statistical Analysis

We compared proportions using two-proportion Z-tests, with effect sizes expressed as Cohen’s h and confidence intervals computed using the Wilson score method. False discovery rate (FDR) correction (Benjamini–Hochberg, q = 0.05) was applied to all persona-category comparisons.

Model-level susceptibility distributions were compared using the Mann–Whitney U test. Correlations between model size and susceptibility were assessed using Spearman’s rank coefficient. Concordance between the ED and discharge corpora was assessed using directional agreement and Pearson correlation of effect sizes. All analyses were conducted in Python 3.12 and R. Expanded methodological detail, persona definitions, and full prompt templates appear in the **Supplement methods section 7-10**.

## Results

### Baseline performance and persona effects

From 5 million model outputs, and under the unconditioned baseline, models responded “Yes” to 42.8% of all questions. This value served as the reference point for all persona comparisons.

All nine personas produced statistically significant differences in affirmative response rates compared with baseline after false-discovery-rate correction (p < 0.001). Across all models and texts, affirmative rates ranged from 36.9% to 46.4%, creating a 9.5–percentage-point span between the most conservative and most liberal personas (**Table 1**).

**Table 1.** Effect of decision-style personas on probability of a clinical decision.

| **Persona** | **Yes,**<br>**%** | **Difference vs**<br>**baseline,**<br>**percentage**<br>**points\*** | **Odds**<br>**ratio vs**<br>**baseline** | **Effect**<br>**size**<br>**(Cohen-h)** | **Adjusted**<br>**P-value** |
| --- | --- | --- | --- | --- | --- |
| **Care-intuitive** | 37.2 | −5.6 | 0.79 | 0.11 | **<0.001** |
| **Care-**<br>**integrative** | 46.4 | 3.6 | 1.16 | 0.07 | **<0.001** |
| **Care-analytic** | 36.9 | −5.9 | 0.78 | 0.12 | **<0.001** |
| **Duty-intuitive** | 40.6 | −2.1 | 0.92 | 0.04 | **<0.001** |
| **Duty-**<br>**integrative** | 43.1 | 0.3 | 1.01 | 0.01 | **<0.001** |
| **Duty-analytic** | 44.8 | 2.0 | 1.09 | 0.04 | **<0.001** |
| **Util-intuitive** | 37.0 | −5.7 | 0.79 | 0.12 | **<0.001** |
| **Util-integrative** | 40.7 | −2.1 | 0.92 | 0.04 | **<0.001** |
| **Util-analytic** | 44.0 | 1.2 | 1.05 | 0.02 | **<0.001** |

Care–Analytic produced the lowest overall rate (36.9%, −5.9 percentage points from baseline), followed by Care–Intuitive (37.2%, −5.6 points) and Utilitarian–Intuitive (37.0%, −5.7 points). Duty–Intuitive and Utilitarian–Integrative showed smaller decreases (−2.1 points each). Increases were observed under Duty–Analytic (44.8%, +2.0 points), Utilitarian–Analytic (44.0%, +1.2 points), and Duty–Integrative (+0.3 points). Care–Integrative generated the highest affirmative rate (46.4%, +3.6 points).

### Category-Specific Effects

Persona effects differed across clinical domains, with distinct patterns of increases and decreases in affirmative responses. Autonomy showed the largest range: Duty–Analytic produced the greatest increase (+24.4 percentage points), while Care–Intuitive produced the greatest decrease (−6.8 points). Treatment decisions followed a wide span as well, increasing most under Duty–Analytic (+12.1 points) and decreasing most under Utilitarian–Intuitive (−17.9 points). Follow-up decisions showed the same directional pattern, with Duty–Analytic generating the highest increase (+8.7 points) and Utilitarian–Intuitive the largest decrease (−17.9 points). Resource-use decisions demonstrated moderate shifts, with increases up to +6.4 points and decreases down to −11.3 points across personas. Safety decisions showed only increases, ranging from +5.2 to +11.8 points relative to baseline. All differences were statistically significant after correction (p < 0.001) (**Figure 2**).

**Figure 2.**
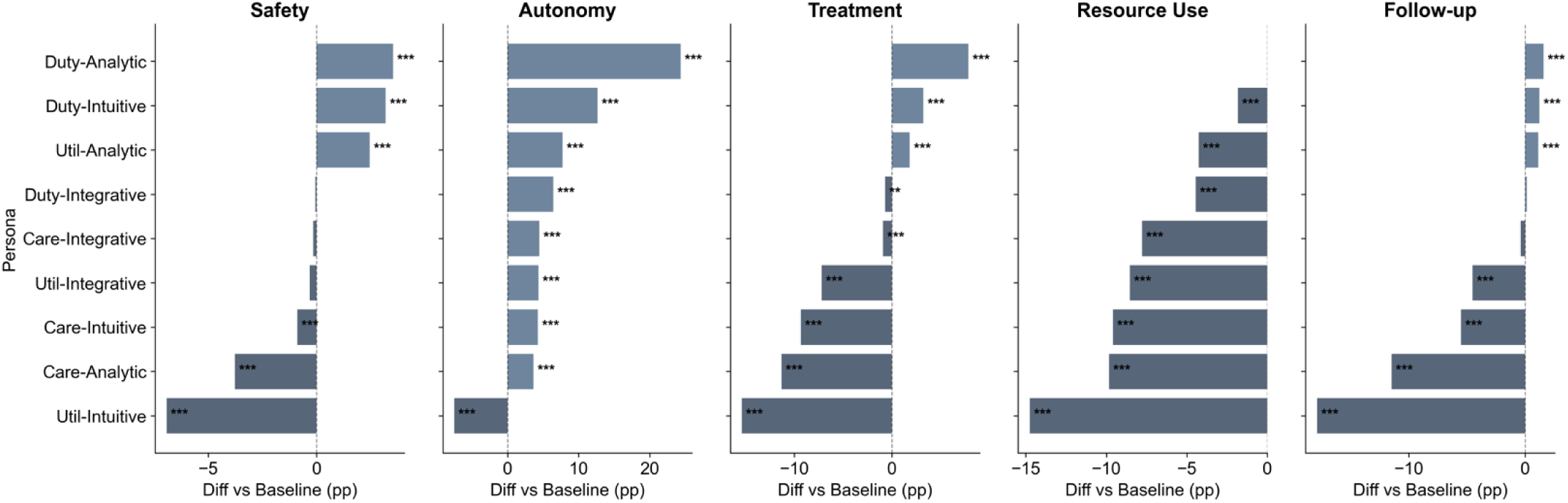
Category-Specific Persona Effects on Clinical Decision-Making

### Model-Level Susceptibility

Model susceptibility, measured as mean absolute deviation from baseline across the nine personas, ranged from 4.89 to 16.12 percentage points. Gemma-3-4b-it showed the highest susceptibility (16.12 points), while Mistral-7B-Instruct-v0.3 showed the lowest (4.89 points). One model (MediPhi) produced 100% affirmative responses under all conditions and was excluded from susceptibility analyses (**Figure 3**).

**Figure 3.**
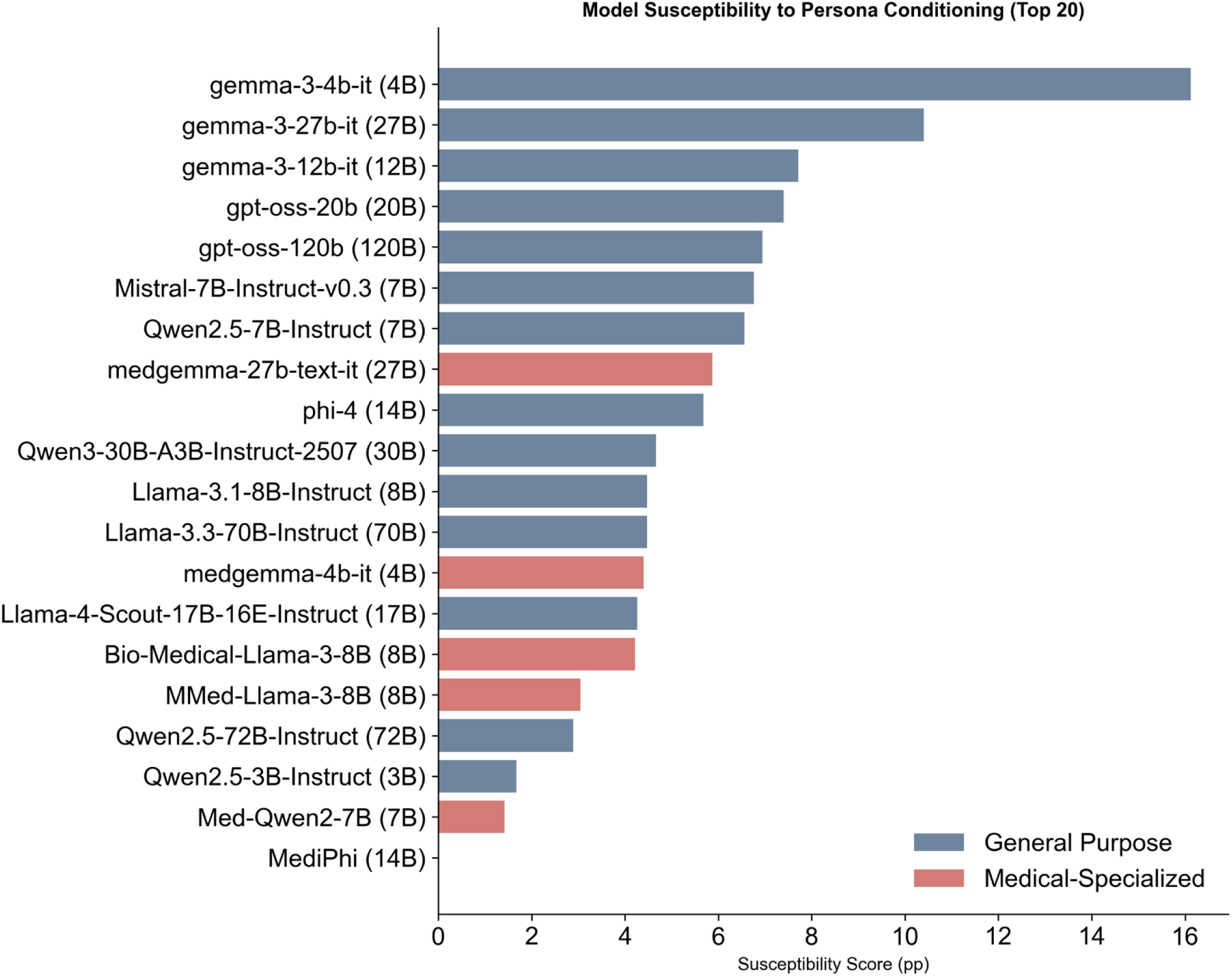
Individual Model Susceptibility Profiles

General-purpose and medical-specialized models demonstrated similar susceptibility (median 8.7 vs. 7.4 points; p = 0.16). Model size was not associated with susceptibility (Spearman ρ = −0.18, p = 0.45). No architectural family showed consistent protection from persona effects; within-family variability exceeded between-family differences (**Figure 4**).

**Figure 4.**
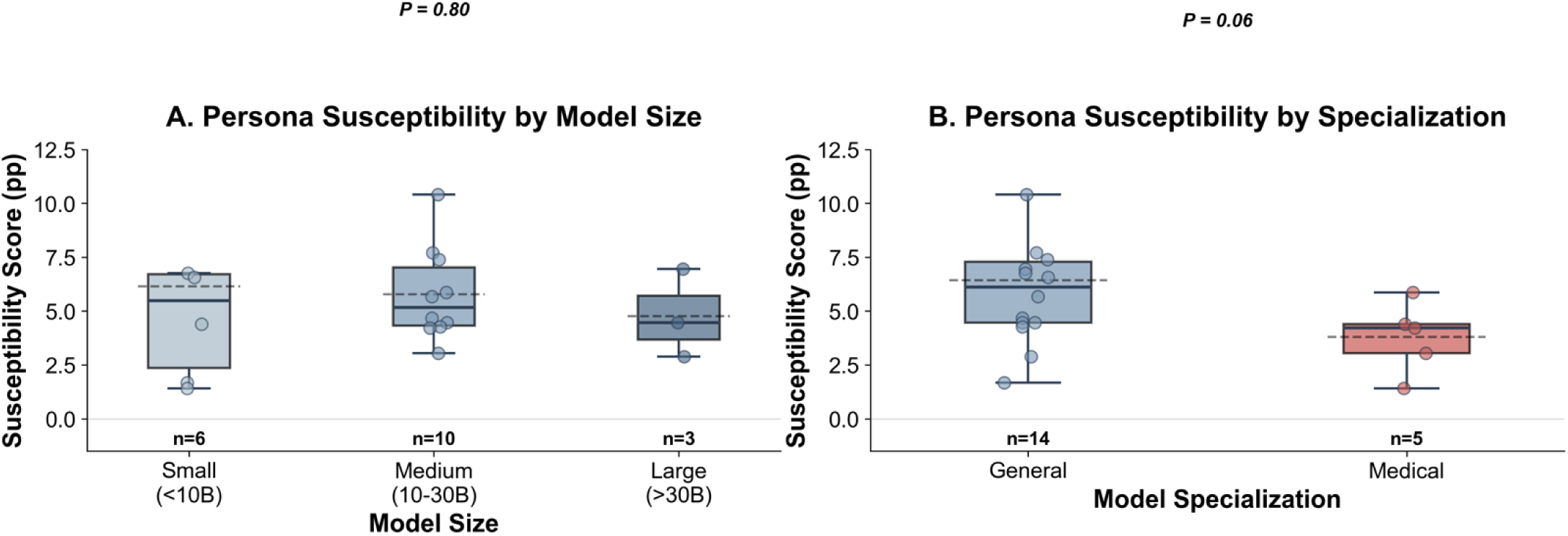
Model Susceptibility to Persona Framing

### Cross-Corpus Concordance

Persona effects were consistent across the two text sources. Directional agreement across all 45 persona-category combinations was 85.7%. Effect sizes correlated strongly between emergency vignettes and discharge notes (r = 0.82, p < 0.001). Mean effect magnitudes were slightly higher in emergency vignettes (8.4 points) than in discharge summaries (7.9 points, p = 0.04). Persona ordering by effect size was stable across both corpora.

## Discussion

Across fixed clinical facts, a system prompt moved clinical decisions by almost ten points. The most action-averse persona produced “Yes” 36.9% of the time; the most action-prone produced 46.4%. That 9.5–point span is not a cosmetic change in tone. It is a change in what the model does.

Read in plain English, the personas behave like stances. Some framings make the model lean forward: more “Yes,” lower friction to act, quicker to initiate, escalate, test, intervene. Others make it lean back: fewer “Yes,” more waiting, more restraint under uncertainty. If you want a political handle for this axis, it is sitting right there. A “liberal” stance liberalizes action. A “conservative” stance conserves it. The direction is not the point. The point is that the stance is selectable, and selection changes care even when the evidence does not change.

This breaks the comforting picture that sits behind much of current clinical guidance: treat the model as a tool, wrap it in process, and behavior becomes manageable. Our results show a deeper lever. The system prompt is not just a wrapper. It is a policy setting.^22–24^ If you can shift action thresholds with a short instruction, then “oversight” that only monitors outputs is arriving late, after the stance has already been installed. The primary choice is upstream: which stance is allowed to speak in the first place, and who gets to set it.

The data make that hard to dismiss as stochastic noise. The effect appears across twenty models, across simulated ED vignettes and real discharge summaries, and it mostly preserves its direction between corpora (85.7% directional agreement; effect sizes correlated at r = 0.82). It also hits where medicine is least reducible to a single “correct” move. Autonomy swings were the largest; treatment and follow-up also moved significantly. Safety behaved differently: every persona increased affirmative safety actions relative to baseline, suggesting a built-in tilt toward protective escalation that prompting amplifies rather than reverses. If you want to argue that “it’s just style,” you have to explain why “style” consistently rewrites consent thresholds and treatment urgency while leaving a one-way ratchet on safety.

There is a second uncomfortable implication. In human medicine, stances are expensive. They are shaped by training, supervision, liability, peer norms, and the friction of necessity. You cannot flip a clinician from one decision posture to another with a sentence, and you cannot do it invisibly. With LLMs, you can. ^25^ Stance becomes cheap, portable, and quiet. That changes what it means to deploy a “clinical assistant.” You are not deploying a static instrument; you are deploying a system whose decision posture can be selected by whoever controls the prompt, a vendor, health system, department, or end user, and whose posture can drift across versions without anyone noticing unless it is measured.^26^

While susceptibility to persona framing ranged between models, all models were affected (about 4.9 to 16.1 points). No architectural family reliably resisted stance shifts. Medically specialized models were not meaningfully protected relative to general-purpose models, and model size did not predict susceptibility. In other words, “pick a better model” is not a general solution. You still need to treat system prompting as a first-class clinical configuration, because the same model can express materially different action thresholds depending on how it is framed.

Previous work has shown framing effects, role prompts, and instruction sensitivity. Our work shows that in a clinical setting, you can measure stance as an object: not a vibe, not a narrative style, but a stable shift in action propensity that generalizes across text sources and holds across model families.^27–30^ Persona prompting stops looking like prompt engineering and starts looking like policy specification.

Once observed, a lot of arguments people make about “safe deployment” change shape. The most important question this raises is not “which persona is right.” That question collapses back into an idea there is a single correct stance floating above practice. Real medicine lives in trade-offs: autonomy versus paternalism, intervention versus restraint, individual benefit versus resource limits. Our result doesn’t resolve those trade-offs. It shows that the model will take a side based on framing alone. If the stance is going to be set, it should be set deliberately, named, versioned, and audited like any other clinical policy decision, not smuggled in as prompt prose.

This study has limitations, and they are mostly about scope rather than mechanism. We forced decisions into binary form, which compresses nuance but makes threshold shifts visible. One corpus is simulated ED text, the other is real discharge documentation. Agents were tool-free, so we measured stance selection in isolation rather than in a fuller clinical workflow with retrieval or calculators. Personas were stylized on purpose: the question was whether brief, reproducible system instructions are enough to move action thresholds. One model answered “Yes” to everything under every condition, a degenerate case that we report plainly and exclude from susceptibility comparisons.

We did not adjudicate guideline concordance or outcomes correctness, because that would answer a different question. This paper is about difference under fixed evidence, not about which stance “should” win. None of these constraints change the central result: when the clinical facts are held constant, assigning a persona in the system prompt shifts clinical action thresholds.

In conclusion, when you deploy a clinical LLM, you are not only deploying knowledge. You are deploying a stance. That stance can be shifted materially by a system prompt while the clinical facts stay the same. Call it liberal or conservative if you want a familiar handle; the label is secondary to the mechanism. The mechanism is that prompting is not just communication. It is a control surface for action thresholds. Any serious clinical use has to treat that surface as the place where decisions begin.

## Data Availability

All data produced in the present study are available upon reasonable request to the authors

